# Modelling optimal vaccination strategies against Covid-19 in a context of Gamma variant predominance in Brazil

**DOI:** 10.1101/2021.11.19.21266590

**Authors:** Leonardo Souto Ferreira, Gabriel Berg de Almeida, Marcelo Eduardo Borges, Lorena Mendes Simon, Silas Poloni, Ângela Maria Bagattini, Michelle Quarti Machado da Rosa, José Alexandre Felizola Diniz Filho, Ricardo de Souza Kuchenbecker, Suzi Alves Camey, Roberto André Kraenkel, Renato Mendes Coutinho, Cristiana Maria Toscano

## Abstract

Brazil experienced moments of collapse in its health system throughout 2021, driven by a timid initial vaccination strategy against Covid-19, combined with the emergence of variants of interest (VOC). Mathematical modelling has been used to subsidize decision-makers in public health planning. Considering the vaccine products available, effectiveness estimates, the emergence of Gamma as the predominant VOC circulating in 2021, and national estimated doses available for the next several months, we developed a Markov-chain mathematical modelling approach to evaluate optimal strategies for Covid-19 vaccination in Brazil in terms of Covid deaths averted. Our main findings are that in order to reach higher vaccination impact in Brazil, Covid-19 immunization strategies should include recovering vaccination coverage rates in high-risk groups reaching higher coverage; expanding vaccination to younger age groups should be considered only after ensuring at least 80% coverage in older age groups; reducing the interval between doses of AZD1222 from 12 to 8 weeks. We also demonstrate that the latter is only feasible if accompanied by an increase in vaccine supply of at least 50% in the next six month period.

## Introduction

To date, Brazil has reached over 600,000 deaths due to Covid-19, ranking second in the world for the absolute number of Covid-19 deaths (1). The first Covid-19 case was identified in the country in March 2020, and the first wave of the disease reached its first peak in July. By October 2020, when more than 5 million confirmed cases and at least 150.000 deaths had been registered in Brazil (2), the first peak of the disease had been overcome. The downward trend that lasted 16 weeks ceased in late November 2020, when a new rise in cases and deaths was observed (3). This coincides with the detection of the variants of concern (VOC) in the country, challenging Brazil’s health systems and the government’s public response. As the fifth-largest world’s country, Brazil presented multiple and different epidemic curves according to the transmission of SARS-CoV-2 between regions in 2020, progressively reaching countryside smaller cities and developing a national synchronization process between 2020 and 2021 where outbreaks caused by VOC, such as Delta and Gamma, were important drivers of sustainable transmission. The Gamma variant (P1 lineage or GR/501Y.V3), first identified in Brazil (3), spread throughout the country before the national vaccination campaign, resulting in numerous surges throughout the territory. In the tenth epidemiological week (EW) of 2021, there was an impressive number of 40,797 hospital admissions, corresponding to an increase of 192% compared with the worst week of the epidemic in 2020 (EW 28) (4).

Covid-19 vaccination rollout started on EW three of 2021 and followed a prioritization framework proposed by WHO (5), initially targeting health professionals, older adults, and people with comorbidities, in this order. Vaccination started slowly and restrictively in its first weeks; the daily-applied doses 7-day rolling average was around 200 thousand doses a day (6). From January to March, vaccination was jeopardized in different moments by limited vaccine supply. Only after April, the daily number of administered vaccines reached 600 to 700 thousand (6). The current Covid-19 vaccines deployed in Brazil are AZD1222 (AstraZeneca/Oxford/Fiocruz), a two-dose adenovirus vaccine, currently administered in a 12-week dose-interval; CoronaVac (Sinovac/Butantan), a two-dose inactivated virus vaccine, using a 4-week dose-interval; BNT162b2 (Pfizer/BioNTech), a two-dose mRNA vaccine, currently administered in a 12-week dose-interval; and Ad26.COV2.S (Janssen) a single dose viral vector vaccine.

National vaccine rollout and immunization strategy were determined in a context of high first-dose effectiveness for the original virus, demonstrated in phase 3 clinical trials (7–9). With the advent of emerging VOC and amid an intense second wave of the pandemic, on EW 34 in 2021 the Brazilian Ministry of Health (MoH) opted to extend the dosing interval of 2 of the three available vaccines at the time (AZD1222 and BNT162b2 vaccines) to 12 weeks, following WHO recommendations (10,11). The objective of such an initiative was to increase the number of vaccinated high-risk group individuals with at least one dose and thus increase the population-level impact in a context of a limited supply of available vaccines.

The Brazilian National Immunization Programme (PNI) was created in 1973 at the end of the smallpox eradication initiative. It represents a robust public health programme offering, free of charge, vaccines incorporated into the routine immunization schedule to all populations through its publicly-funded Universal Health Care System (SUS) (12). Managed by the Federal Government, together with States and Municipalities, decentralized and with good capillarity, achieved by more than 36 thousand rooms in 5,570 Brazilian cities, the PNI has historically been able to deliver massive amounts of vaccines in immunization campaigns and get to hard-to-reach populations. In 2010, for example, in response to the Influenza pandemic, the MoH managed to administer almost 90 million doses of Influenza vaccines in only five months (13).

Differently from what would be expected in a country with this record on immunization activities, the National Immunization Campaign for Covid-19 in Brazil has faced several new obstacles. These included lack of national coordination and support to evidence-based decision making, inconsistent vaccine supply and availability over time, limited social communication strategies, and widespread social networks misinformation about vaccine safety and efficacy. In addition, the anti-vax movement in the country has been gaining strength throughout the SARS-CoV-2 dissemination process. From the operational point of view, the Covid-19 vaccination rollout was fragmented, with different vaccination strategies being adopted as a decentralized process involving state and municipal levels. In a country of continental dimensions experiencing different regional and local pandemic waves, the lack of communication and an integrated response brought additional challenges to vaccination implementation strategies. As such, vaccines were progressively made available to adults without comorbidities, by age, irrespective of vaccine coverages in priority risk populations. One critical point is that, in this age-based strategy, a minimum coverage goal set by age group was not established before the vaccine was made available for younger groups. This progression was based on a temporal criterion (for example, a one-week period designated for each age group), varying by locality.

Considering the vaccine products available, effectiveness estimates, the emergence of Gamma as the predominant VOC circulating in 2021, and national estimated doses available for the next several months, we developed mathematical modelling approaches to evaluate optimal strategies for COVID-19 vaccination in Brazil. This work aims to provide immediate answers to pressing questions and a proposed framework for future analyses to provide decision-makers with modelling evidence to support the decision-making process at a national level regarding best strategies to minimize the burden of Covid-19 deaths in Brazil.

We specifically aim to address three programmatic questions: what should be the vaccination coverage of an age group before starting vaccination in a younger group? What should be the ideal interval between doses of the AZD1222 vaccine that would result in the greatest impact (considering intervals between 8 and 12 weeks), assuming a sufficient supply of vaccines? Finally, what is the minimum amount of vaccines made available over time that will allow the implementation of the optimal interval between doses?

## Methods

### Data sources

We use the current number of vaccinated individuals from the Information System of the National Programme on Vaccination (SI-PNI) from Brazil (2021-08-09 dataset) (6). The dataset contains anonymized information of each vaccinated individual in Brazil, first or second dose identification, the name of the vaccine, date of inoculation, and the individual’s age.

The model considers the current vaccines implemented in Brazil: AZD1222 (AstraZeneca/Oxford/Fiocruz), CoronaVac (Sinovac/Butantan) and BNT162b2 (Pfizer/BioNTech). Since our goal is to estimate optimal strategies considering the interval between vaccine doses, we have not included the Ad26.COV2.S vaccine in the model. Also, the minor frequency of individuals that received or will receive this vaccine (4.5 million people out of 211 million people, representing less than 2% of the population) means that this should not affect our conclusions.

We use the number of doses procured by the Brazilian Ministry of Health and anticipated to be delivered until the end of 2021 as input of the model (6). Since the number of doses is projected by month or quarter, we assume a constant production rate in each time interval (See Supplementary material). With this information, we estimate the number of individuals with two doses per vaccine and age group. We also estimate the number of individuals with only one dose per vaccine, age group, and time since first-dose vaccination. This data feeds the mathematical model described in the next section.

### Model

To estimate the number of deaths averted by different vaccination strategies, we developed a discrete-time Markov Chain model with a constant probability of infection (14). To avoid modelling stochastic dynamics, we use the mean-field approximation where the probability of infection turns to a proportion of susceptible individuals infected per day.

The structure of the model is the same as an extended SEIR model (15), accounting also for asymptomatic, hospitalized, and deceased individuals, thus being named SEAIRHD (16). The model is age-structured in 10-year groups. This structure is replicated for each dose (first and second dose) of each vaccine (AZD1222, CoronaVac, and BNT162b2), and the vaccines are modelled simultaneously (see Figure 1).

**Figure 1.**
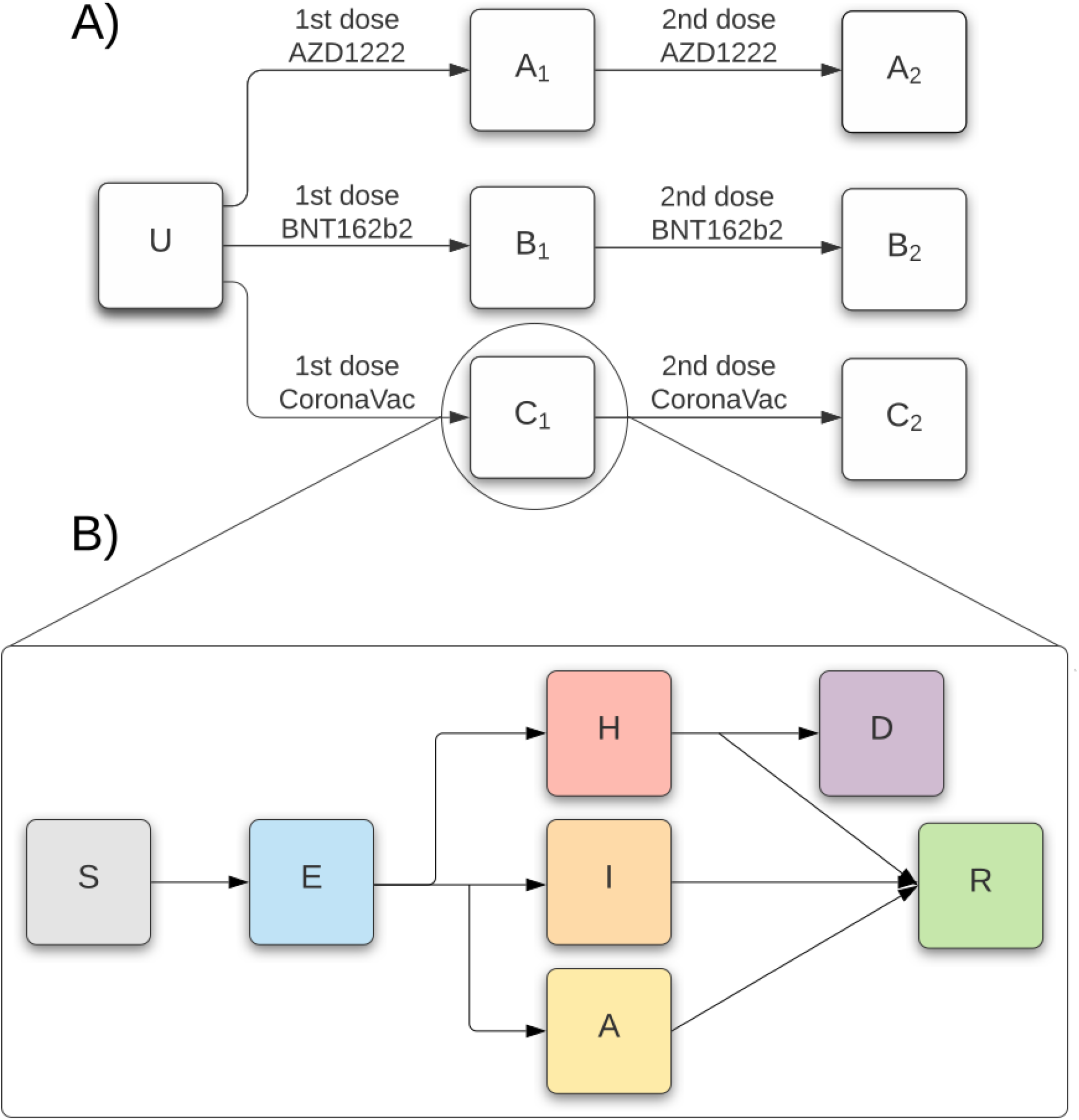
Structure of the mathematical model. The first diagram (**A**) describes the vaccination pathways. U accounts for unvaccinated individuals; A, B, and C accounts for individuals vaccinated with AZD1222, BNT162b2, and CoronaVac, respectively. The subscripts account for the first (1) or second (2) dose. The second diagram (**B**) describes the infection pathways. A susceptible (S) individual is transferred to a pre-symptomatic (or exposed, E) compartment after infection. After the incubation period, the individual evolves to an asymptomatic (A), mild (I), or severe (H) infection. The individual eventually recovers (R) or dies (D), if severe.

To estimate the number of doses of vaccine that should be allocated for the first or second dose, we use a modified version of the optimization model developed by (16) that accounts for varying production (or deployment) rates and also previously vaccinated individuals with only one dose (see further details on the SM). This optimization model calculates the number of first or second doses used by day, given production rate and the interval between doses, minimizing the number of doses that should be kept in stock while guaranteeing that individuals receive the second dose when recommended.

We assumed a single model for the entirety of Brazil. We also assume that after initially targeting high-risk populations, vaccination rollout followed an age-prioritization. We considered the number of administered doses by vaccine, over time, as being proportional to the total number of doses of each vaccine made available at the time of vaccination. We considered that the vaccination rate in each age group is proportional to the unvaccinated population in this group (more details in Supplementary material).

We limited the analysis of optimal time between doses (and the required supplied doses) only to AZD1222 for the following reasons: 1) there is no evidence to support the use of CoronaVac vaccine in a longer interval than the recommended four weeks. 2) At the time of our modelling, there was no data on BNT162b2 effectiveness against the Gamma variant. In addition, differently than AZD1222 and CoronaVac, this BNT162b2 vaccine is imported, and thus there is no possibility of expanding production and distribution capacity.

### Model Parameters

We assume four scenarios of the probability of Covid-19 infection: very low, low, medium, and high (with numerical values 0.0001, 0.0025, 0.0050, and 0.0100, respectively).

Since Brazil does not have systematic serological inquiries, the exact prevalence of Covid-19 in the population is unknown. We assume that the percentage of recovered individuals at the start of the simulation is drawn from a uniform distribution with a minimum and a maximum of 40% and 60%, respectively. These values vary by each seroprevalence study, with different methodologies and study sites (17,18). We then use a rather wide guess in our model to include most of the possible scenarios.

For the effectiveness of the vaccines, we used recently published data from Cerqueira-Silva et al., who studied AZD1222 and CoronaVac in a retrospective cohort of more than 60 million vaccinated individuals in Brazil in a Gamma predominance scenario. The vaccine effectiveness (VE) results are presented stratified by age, by the outcome, and whether the vaccination scheme is complete (with two doses) or incomplete (only one dose). We assumed that CoronaVac effectiveness against SARS-CoV-2 infection was zero, as this is an inactivated vaccine and considering available evidence from the literature (19). Concerning AZD1222 and BNT162b2, the best available results of VE against infection were used in this work. VE parameters for CoronaVac, AZD1222, and BNT162b2 are presented in tables 1, 2, and 3, respectively. We draw the values of the effectiveness of the vaccines from the Beta distributions that obey the confidence interval of each estimate. These values are stored, and each scenario of the probability of infection and vaccination strategy are simulated with the same values of seroprevalence and effectiveness, thus ensuring comparability of strategies. Each scenario is then run using 500 combinations of values for each result shown in the next section.

**Table 1.**
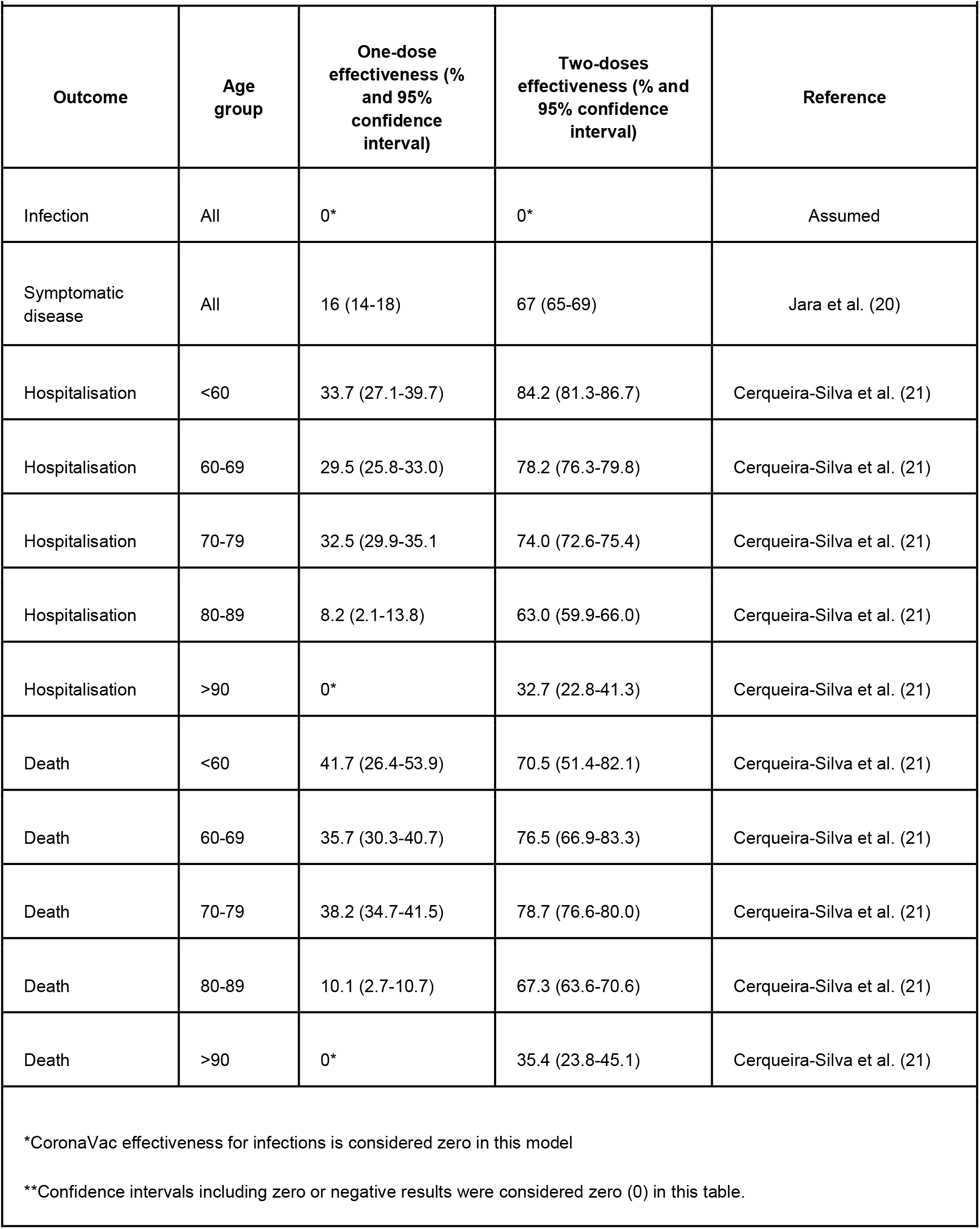
CoronaVac vaccine effectiveness against Gamma Variant of Concern, by outcome, age group, and vaccine dose.

| Outcome | Age group | One-dose effectiveness (% and 95% confidence interval) | Two-doses effectiveness (% and 95% confidence interval) | Reference |
| --- | --- | --- | --- | --- |
| Infection | All | 0* | 0* | Assumed |
| Symptomatic disease | All | 16 (14-18) | 67 (65-69) | Jara et al. (20) |
| Hospitalisation | <60 | 33.7 (27.1-39.7) | 84.2 (81.3-86.7) | Cerqueira-Silva et al. (21) |
| Hospitalisation | 60-69 | 29.5 (25.8-33.0) | 78.2 (76.3-79.8) | Cerqueira-Silva et al. (21) |
| Hospitalisation | 70-79 | 32.5 (29.9-35.1) | 74.0 (72.6-75.4) | Cerqueira-Silva et al. (21) |
| Hospitalisation | 80-89 | 8.2 (2.1-13.8) | 63.0 (59.9-66.0) | Cerqueira-Silva et al. (21) |
| Hospitalisation | >90 | 0* | 32.7 (22.8-41.3) | Cerqueira-Silva et al. (21) |
| Death | <60 | 41.7 (26.4-53.9) | 70.5 (51.4-82.1) | Cerqueira-Silva et al. (21) |
| Death | 60-69 | 35.7 (30.3-40.7) | 76.5 (66.9-83.3) | Cerqueira-Silva et al. (21) |
| Death | 70-79 | 38.2 (34.7-41.5) | 78.7 (76.6-80.0) | Cerqueira-Silva et al. (21) |
| Death | 80-89 | 10.1 (2.7-10.7) | 67.3 (63.6-70.6) | Cerqueira-Silva et al. (21) |
| Death | >90 | 0* | 35.4 (23.8-45.1) | Cerqueira-Silva et al. (21) |

\*CoronaVac effectiveness for infections is considered zero in this model
\*\*Confidence intervals including zero or negative results were considered zero (0) in this table.

**Table 2.**
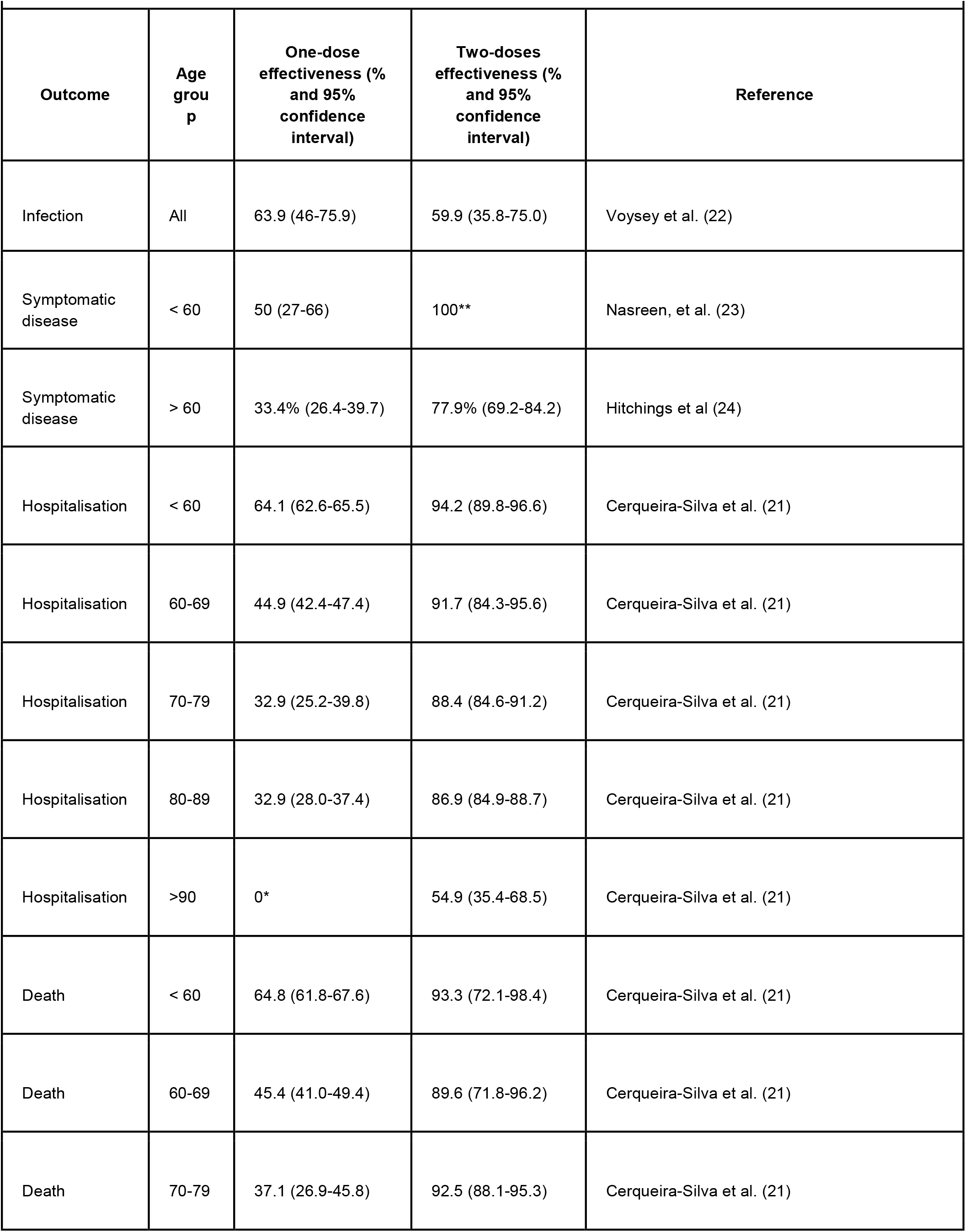
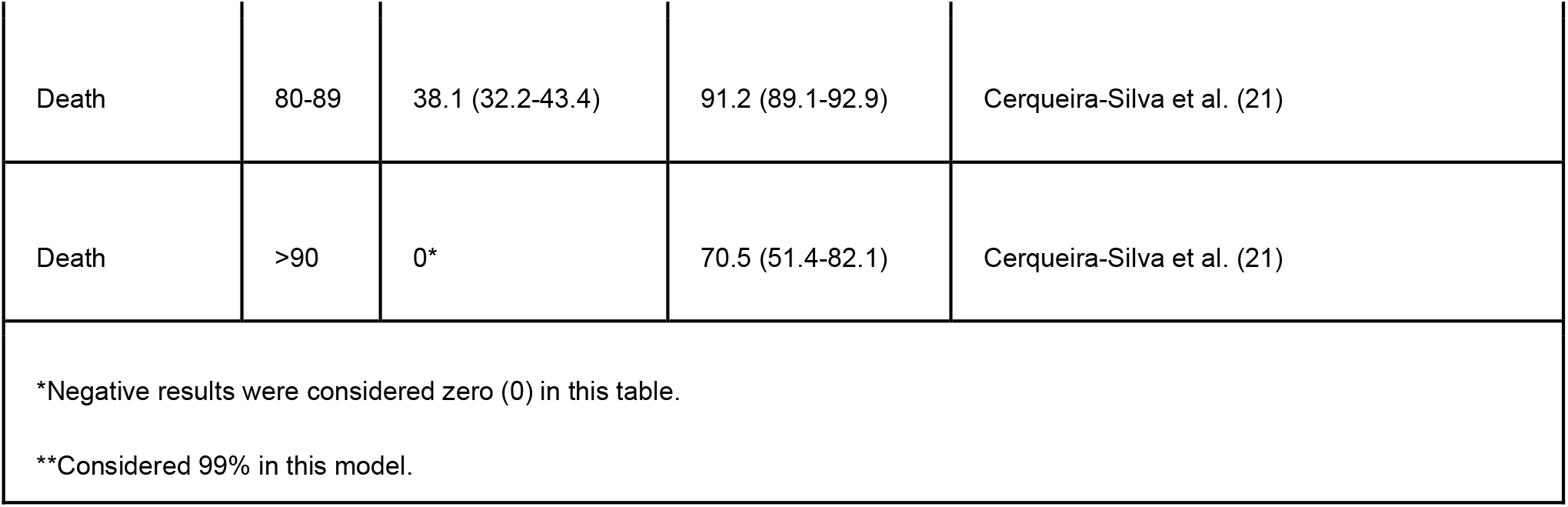
AZD1222 Vaccine Effectiveness against Gamma Variant of Concern, by outcome, age-group, and vaccine dose.

**Table 3.** BNT162b2 vaccine effectiveness against Gamma Variant of Concern, by outcome, age-group and vaccine dose.

|  |  |  |  |  |
| --- | --- | --- | --- | --- |
| **Outcome** | **Age group** | **One-dose effectiveness (% and 95% confidence interval)** | **Two-doses effectiveness (% and 95% confidence interval)** | **Reference** |
| Infection | All | 60 (53-66) | 92 (88-95) | Dagan et al. (25) |
| Symptomatic disease | All | 65 (56-71) | 85 (70-93) | Nasreen et al. (23) |
| Hospitalisation/Deaths | All | 83 (75-88) | 98 (82-100) | Nasreen et al. (23) |

## Results

### What should be the vaccination coverage of an age group before starting vaccination in a younger group?

We calculated the excess deaths caused by starting vaccination in a younger age group after a coverage threshold compared to the strategy of only starting to vaccinate a younger age group after fully vaccinating the older age group. As shown in Figure 2, the lower the vaccination coverage reached in older age groups, the greater the estimated excess of deaths, regardless of the probability of infection. However, we find that the magnitude of the impact is smaller when the probability of infection is lower. Furthermore, it can be seen that at least 90% vaccination coverage is necessary for a minimal excess of deaths to be reached (varying from 10.57 (95%CI: 10.60-13.43) to 620.21 (95%CI: 617.35-774.56) depending on the probability of infection). Nevertheless, starting vaccination in a younger age group when at least 80% vaccination coverage had been reached in older age groups resulted in a third of excess Covid-19 deaths compared to starting vaccination when only 40% coverage had been reached, as shown in Figure 2 below. The estimates for all values of threshold are tabulated in the SM.

**Figure 2.**
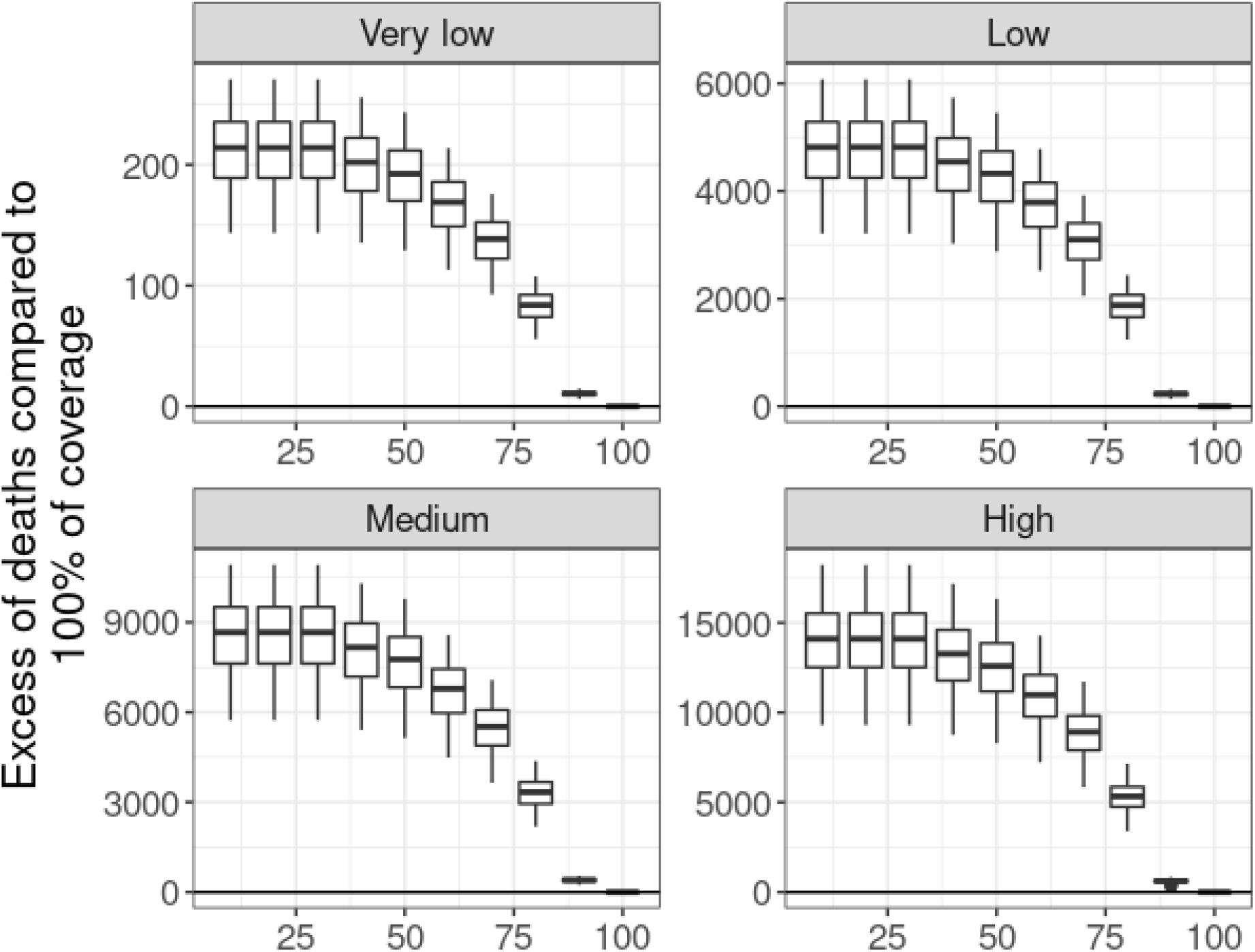
Covid-19 number of excess deaths (median and interquartile ranges) (y-axis) by minimum 2-dose coverage threshold (x-axis) reached before initiating vaccination of younger age groups, stratified by the probability of infection (panels), given by very low, low, medium, and high (with numerical values 0.0001, 0.0025, 0.0050, and 0.0100, respectively) values.

### What should be the ideal interval between doses of the AZD1222 vaccine that would result in the greatest impact (considering intervals between 8 and 12 weeks), assuming a sufficient supply of vaccines?

Although the results show that 90% coverage is the minimum necessary to reduce excess deaths to minimum levels, we consider 80% as a feasible target vaccination threshold. For this threshold, we computed the difference in Covid-19 deaths estimated when considering AZD1222 vaccine schedules with different dose intervals (8, 9, 10, and 11 weeks) compared to the standard currently recommended 12-week interval (Figure 3). We assumed a setting without limitation of AZD1222 vaccine doses, i.e., we ran the model assuming a number of doses up to ten times higher than the currently available and projected doses of AZD1222. We found that the lower dose-interval of 8 weeks leads to a greater reduction in the number of Covid-19 deaths, varying from 99.56 (95%CI: 64.17-133.82) deaths averted in the lower probability of infection scenario to 9,126.66 (95%CI: 5,941.56-12,298.09) in a scenario of a high probability of infection. All estimates are tabulated in the SM.

**Figure 3.**
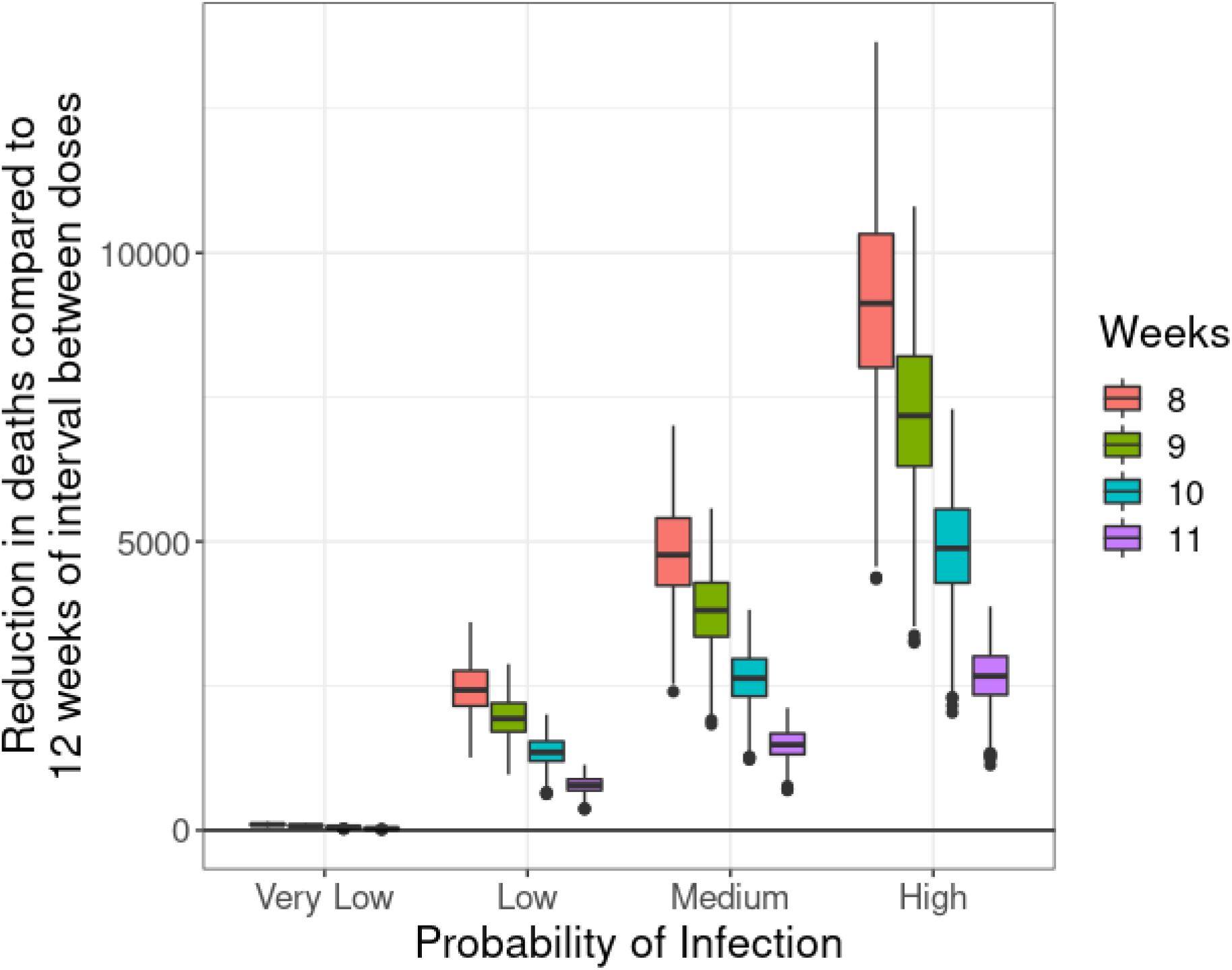
Covid-19 deaths averted (median and interquartile ranges) (y-axis) by different dose intervals (colour) compared to 12 weeks, stratified by the probability of infection (x-axis), under the scenario of no limitation of AZD1222 vaccine supply. The probabilities of infection are given by very low, low, medium, and high (with numerical values 0.0001, 0.0025, 0.0050, and 0.0100, respectively) values.

This result leads to whether the strategy of using a lower dose-interval for AZD1222 would be effective considering the currently projected vaccine supply until the end of 2021. When considering the scenario of the currently projected AZD1222 vaccine supply, we see that the reduction in deaths is negligible, less than one death regardless of the probability of infection (Figure 4). This, in turn, leads to the next question which this modelling addresses.

**Figure 4.**
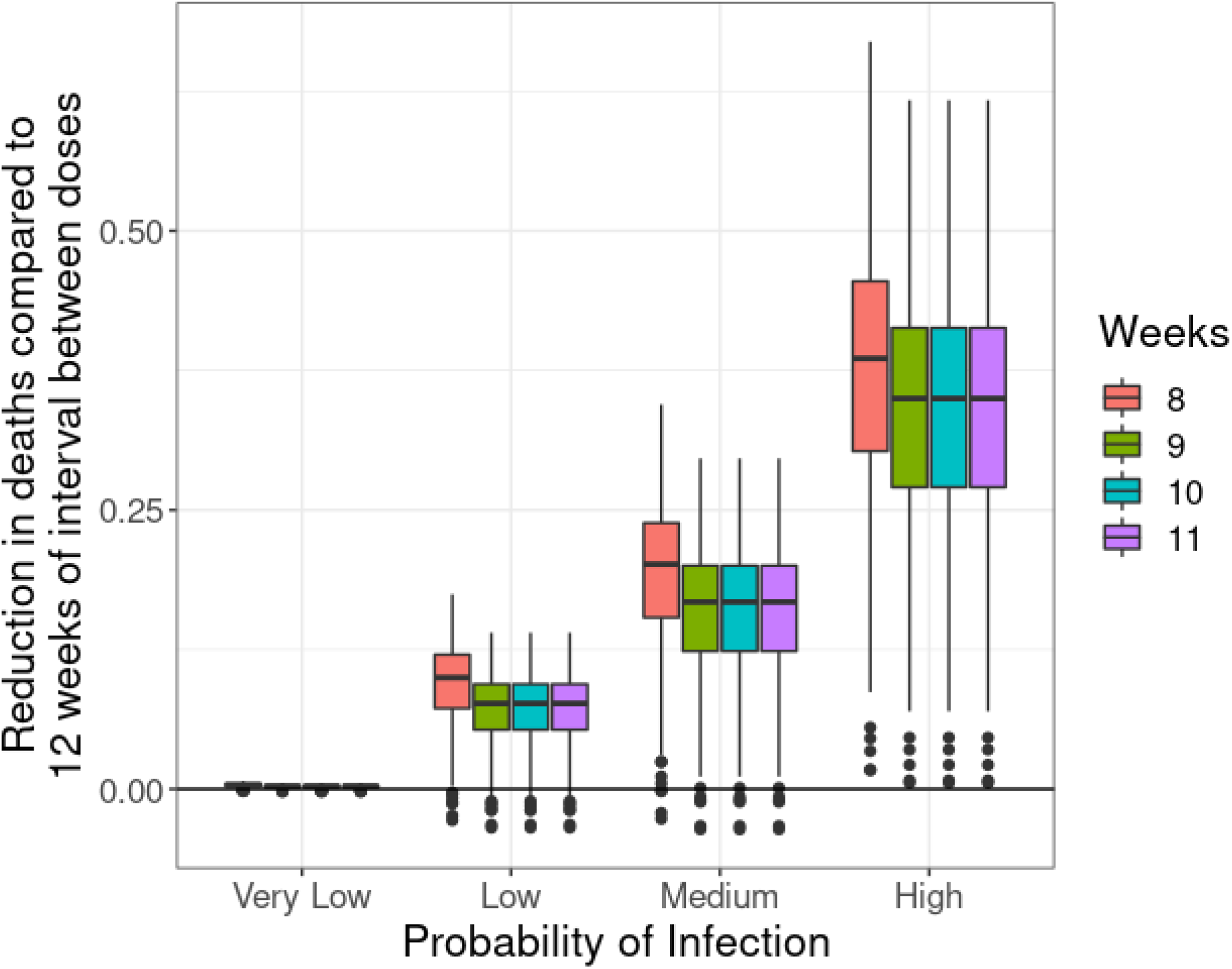
Reduction in deaths (in median percentage and interquartile ranges) (y-axis) by different dose-intervals (colour) compared to 12 weeks, stratified by the probability of infection (x-axis), under the scenario of the currently projected AZD1222 vaccine supply. The probabilities of infection are given by very low, low, medium, and high (with numerical values 0.0001, 0.0025, 0.0050, and 0.0100, respectively) values.

### What is the minimum amount of vaccines made available over time that will allow the implementation of the optimal interval between doses?

Considering the different dose-intervals for the AZD1222 vaccine, we estimate that the number of vaccine doses administered needs to be increased by at least 50% to avoid supply bottlenecks and allow for an impact on Covid-19 deaths reduction be observed (Figure 5). When comparing these estimates to the ones presented in Figure 3, we can observe the different population impact of the strategy when considering a scenario in which enough vaccine doses are available to meet the entire demand (Figure 4) and a scenario in which an insufficient number of vaccine doses is available to meet the demand, thus requiring an increase in vaccine supply (Figure 5). Even with a 100% increase in vaccine supply, the impact of reducing the time between doses to 8 weeks is reduced to at least about one-fifth of the maximum potential impact (reduction from 9,126.66 (95%CI: 5,941.56-12,298.09) to 2018.58 (95%CI: 1,167.27-2,823.14) deaths averted in the scenario of a high probability of infection). A higher increase in vaccine supply was not considered as we assumed this was not a feasible scenario.

**Figure 5.**
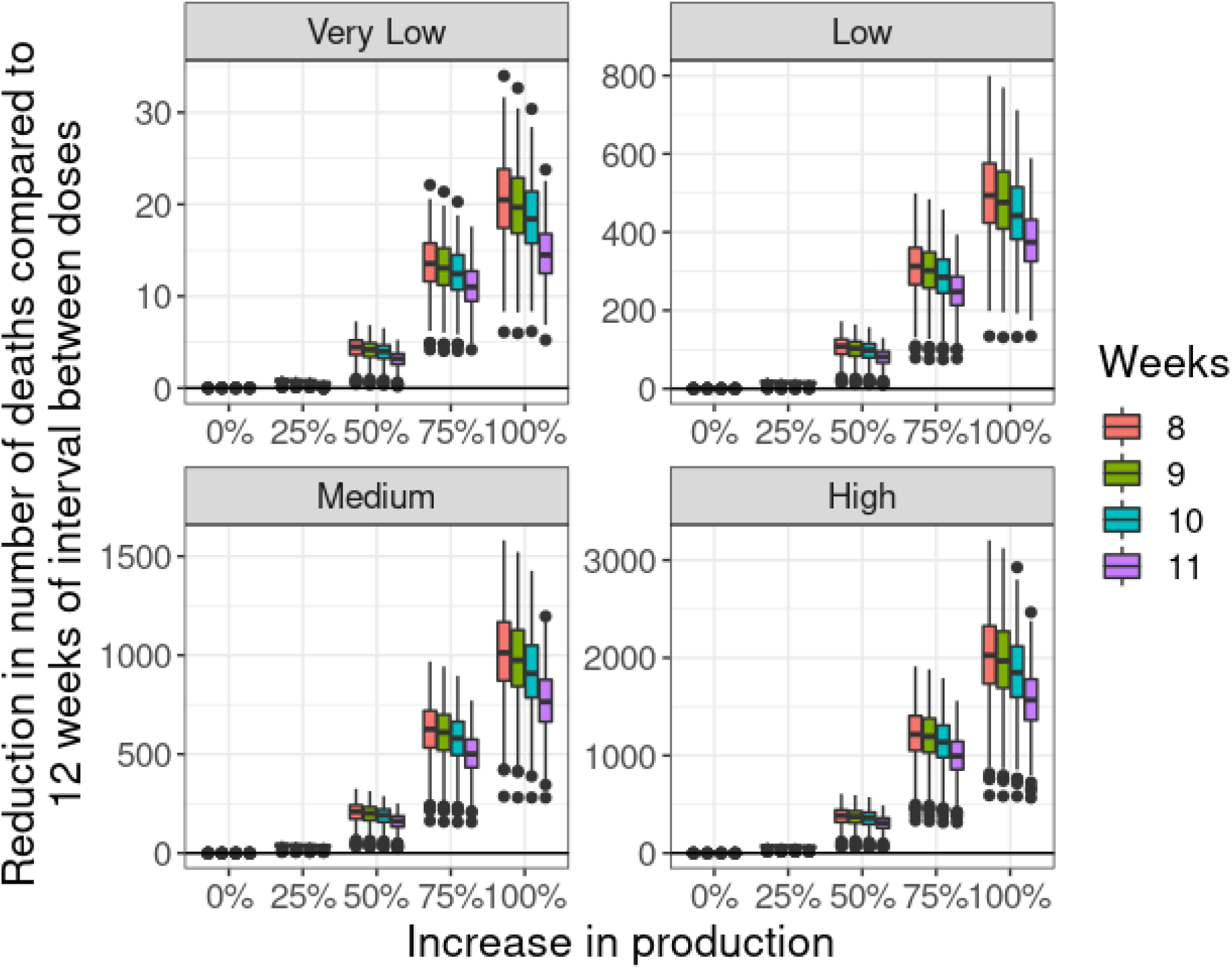
Covid-19 deaths averted (median and interquartile ranges) (y-axis) when considering lower dose-intervals (colour) compared to 12 weeks, by per cent increase in AZD1222 vaccine supply (when compared to current projections) (x-axis), stratified by the probability of infection (panels), given by very low, low, medium, and high (with numerical values 0.0001, 0.0025, 0.0050, and 0.0100, respectively) values.

## Discussion

Experiencing a pandemic requires constant and routine monitoring of the epidemiologic and its evolution over time, and implementation and adjustments of public health policies and disease control measures. Covid-19 challenges Brazil in a context where the Federal Government did not consider the purchase of vaccines as a priority in the fight against the pandemic, in addition to discouraging the use of masks, social distancing and respect for quarantine measures, claiming that these measures would bring enormous losses of economic impact. This, associated with the emergence of new variants of concern (VOC), such as the Gamma variant (26), can represent a major problem. Being flexible and rethinking strategies has been mandatory in a large, plural country without a central plan to fight the epidemic (27).

By implementing vaccination on a large scale, so far with good efficacy data for all vaccines with only one dose for the original virus, Brazil focused on vaccinating the greatest number of people with one dose, ensuring some immunity, and spacing out the second dose for the maximum periods stipulated by the manufacturers. Thus, until July 2021, both the AZD1222 and BNT162b2 vaccines were administered with a 12-week interval between doses. In addition, the eagerness to reach a more significant number of vaccinated individuals has led states and municipalities to expand vaccination including early on younger age groups without defining *a priori* a minimum coverage to be reached in the groups already prioritized, which can also represent a problem.

In the context of scarce vaccine supply, it is crucial to ensure that at-risk individuals are adequately protected against Covid-19. Since our model limits to age-stratified populations, ignoring other groups, for example, pregnant women and immunosuppressed individuals, we can measure the effect of different thresholds of coverage of older individuals before making vaccine doses available to younger individuals. Figure 2 shows that ensuring a good vaccine coverage of older individuals (at least 90% of coverage) reduces the number of deaths considerably, as expected. However, more important than that, using lower coverages (lower than 80%) as a threshold generates a sharp increase in the number of additional deaths compared to vaccinating the whole population of older individuals beforehand. Thus, the first strong recommendation that this article can make to optimize the ongoing vaccination plan in Brazil is that it is necessary to resume efforts to achieve minimum coverage in older age groups, so that, only after a minimum coverage of 80%, we can advance vaccination to younger age groups. This calls for urgent measures from government policy makers, including social mobilization and communication campaigns, in addition to active search of unvaccinated individuals. Assuming that Brazil is perfectly capable of rescuing unvaccinated older people, given the country’s brilliant history concerning vaccination planning and implementation, acceptance by the population, and especially the capillarity of the health system, we can advance on the issues we modelled.

We demonstrate that, considering vaccine effectiveness estimates obtained in a scenario of Gamma variant dominance, reducing the interval between AZD1222 doses from 12 to 8 weeks ensures the highest reduction in Covid-19 deaths (up to 10 thousand deaths in the high transmission scenario), independently of the epidemic situation in Brazil. This study does not investigate smaller intervals between doses, as current guidelines recommend 8-12 weeks between the two doses (11). However, an important question raised by this work is that the optimal impact of changing the interval between doses is not (nearly) achievable by the current vaccine supply (i.e., the projected number of doses to be distributed in Brazil in the next 6 months), as a high number of individuals are left unprotected during the course of the epidemic. Changing the recommendation in the current scenario, with the number of doses currently available, would not be interesting, as it would lead to a bottleneck in vaccine supply with potential damage to the immunization program.

To achieve a noticeable impact of changing the interval between doses of AZD1222, we see that the increase vaccine supply, in terms of number of available doses should be in the order of, at least, 50%, independently of the transmission level of the epidemic. This might be achievable as Brazil’s AZD1222 producer Fiocruz-Biomanguinhos is upgrading its factory from filling doses to in loco production of active pharmaceutical ingredients. This would, in principle, enable the production of four monthly lots of doses instead of the current three lots. Ideally, increasing vaccine availability by 100%, would result in an ideal scenario in terms of vaccination impact. We did not model an increase beyond that level, as we assumed it to be unrealistic. Thus, one more strong recommendation based on mathematical modelling can be made: decreasing the interval between doses of AZ vaccines from 12 to 8 weeks can be highly beneficial, in a scenario of 100% increase in vaccine availability by the end of the year 2021.

This recommendation is important to reinforce the need for increasing and sustaining local vaccine availability and demonstrate that the country cannot let down its guard in the fight against the pandemic. The upsurge in cases of covid-19, which ran from late 2020 to mid-2021, now appears to be showing signs of fading, despite the more recent introduction of the Delta VOC in the country. However, this is observed in the context of a significant proportion of the population either vaccinated or previously infected. Relaxing social distancing measures or slowing down vaccination activities, at this time, may result in increased infection transmission, ultimately leading to, once again, increase in cases and saturation of the healthcare system.

Regarding the Delta VOC, recently introduced and now predominant in Brazil, which was not modelled in this work, recent evidence indicates that effectiveness of one dose of AZD1222 is reduced when compared to 1-dose effectiveness against the Gamma variant. In contrast, the effectiveness post-second dose against severe disease maintains levels comparable to the Alpha variant (23). Thus, one would assume that the same vaccination strategies identified as optimal should be used to contain the disease caused by the Delta variant, i.e., reinforcing vaccination coverage of 60+ age groups to at least 80%, reducing the interval between doses of AZD1222 from 12 to 8 weeks and upgrading the production capabilities to at least 100%.

Mathematical modelling has been extensively used to assist policymakers during the Covid-19 pandemic. The range of scenarios studied includes, but is not limited to, school reopening (28), the effects of lockdown (29), and, of course, vaccination strategies. Moore et al. (30) have shown that vaccinating older age groups should be prioritized to minimize the number of future deaths or years of life lost in the UK. The same results were found by Bubar et al. (31) when considering the number of deaths as the outcome. However, under a highly effective vaccine against infection scenario, vaccinating more mobile age groups reduced the greatest number of infections in the population. Some agent-based models have been used to assess the effects of delaying the second dose of mRNA-based vaccines, showing that under the VE results against Alpha variant, delaying up to 12 weeks would have positive effects on the number of deaths (32,33).

The results presented here are in agreement with those reported by Silva et al. (34) and Ferreira et al (16). Silva et al. (34), using a hypothetical scenario of the number of doses, estimate that with a vaccine that alleviates symptoms with first dose efficacy between 60% and 70%, using an 8-week interval between doses would reduce the most number of ICU admissions. Notice that this is the case when we consider the effectiveness of AZD1222 against hospitalizations/deaths (Table 2). Ferreira et al. (16) considered varying (constant) production rates and have shown that, besides first dose efficacy, knowledge of vaccine production is also an important parameter when considering the optimal interval between doses, whereas the optimal windows between doses are robust under different values of effective reproduction number, a fact confirmed by our modelling, and also supports our choice of fixed scenarios of the probability of infection. This work improves upon these articles by: first, considering variable in time production rates of vaccine, following the procured quantities by the Ministry of Health, instead of hypothetical scenarios. Second, considering all three vaccines simultaneously in the model, optimizing the allocation of doses in an ongoing vaccination programme. Third, using the most up-to-date parameters of effectiveness of vaccines against Variants of Concern.

As the vaccination data is reliable in Brazil and data from recent studies regarding vaccine efficacy for the Gamma variant is available, we were able to implement a Markov Chain mathematical model to assess optimization strategies in the National Immunization Programme (PNI). The main findings of this work are: the Brazilian vaccination programme should recover vaccination rates in at-risk individuals to ensure sufficiently good coverage; starting vaccination in younger age groups should happen after ensuring at least 80% coverage in older age groups; reducing the interval between doses of AZD1222 from 12 to 8 weeks would have the most impact in the number of averted deaths by vaccination, but only if accompanied with an increase of at least 50% of procured vaccine doses by the Ministry of Health.

## Supporting information

Equations of the model, epidemiologic parameters, vaccination parameters, tabulated results.

## Data Availability

The authors state that the database used in the analyses are available as a supplementary file to the paper and can be provided to interested researchers upon reasonable request.

https://github.com/covid19br/paper_markov

## Declaration of interests

The authors have no conflicts of interest.

## Funding and role of the funding source

This study was funded by the Brazilian National Council for Scientific and Technological Development (CNPq) - Process # 402834/2020-8 (request for proposals MCTIC/CNPq/FNDCT/MS/SCTIE/Decit N° 07/2020).

The funding sources played no role in the study design, collection, analysis, or interpretation of the data, writing the report, or deciding to submit the paper for publication.

LSF received a master’s scholarship from Coordination of Superior Level Staff Improvement (CAPES) (finance code 001). GBA received a technological and industrial scholarship from CNPq (grant number 301796/2021-1). MEB received a technological and industrial scholarship from CNPq (grant number 315854/2020-0). LMS received a technological and industrial scholarship from CNPq (grant number 315866/2020-9). SP was supported by São Paulo State Research Support Foundation (FAPESP) (grant number: 2018/24037-4). AMB received a technological and industrial scholarship from CNPq (grant number 402834/2020-8). MQMR received a postdoctoral scholarship from CAPES (grant number 305269/2020-8). RSK has been supported by CNPq (proc. 312378/2019-0). JAFD-F has been supported by the CNPq productivity fellowship and the National Institutes for Science and Technology in Ecology, Evolution and Biodiversity Conservation (INCT-EEC), supported by MCTIC/CNPq (proc. 465610/2014-5) and FAPEG (proc. 201810267000023). RAK has been supported by CNPq (grant number: 311832/2017-2) and FAPESP (contract number: 2016/01343-7). CMT has been supported by the CNPq productivity fellowship and the National Institute of Science and Technology for Health Technology Assessment (IATS) (proc: 465518/2014-1).

## Author contributions

CMT, RAK, JAFD-F, SAC, RSK, and RMC conceptualized the study. All authors reviewed the study design and provided critical comments.

GBA, AMB, LMS, SAC, RSK, MQMR, and CMT reviewed the literature, collected and verified the underlying data, and proposed values and intervals for model parameters. All authors discussed and agreed on model parameters to be considered.

LSF, MEB, SP, and RMC were primarily responsible for model structuring, parametrization, and statistical analyses. All authors participated in the interpretation of the analyses.

LSF and GBA drafted the paper. All authors contributed to revising the paper, the tables, and the figures critically and for important intellectual content.

All authors approved the final submitted version of the paper.

All authors have full access to the data, are accountable for all aspects of the work, and will ensure that questions related to the accuracy or integrity of any part of the work are appropriately investigated and resolved.

## Notes

### Competing Interest Statement

The authors have declared no competing interest.

