## Supplementary material for "Modelling optimal vaccination strategies against Covid-19 in a context of Gamma variant predominance in Brazil": Equations of the model, epidemiologic parameters, vaccination parameters, tabulated results.

Epidemiology and Infection

Supplementary Material

Leonardo Souto Ferreira[1,2], Gabriel Berg de Almeida[3], Marcelo Eduardo Borges[2], Silas Poloni[1,2], Lorena Mendes Simon[4], Angela Maria Bagattini[5], Michelle Quarti Machado da Rosa[5], José Alexandre Felizola Diniz Filho[2,4], Ricardo de Souza Kuchenbecker[6], Suzy Alves Camey[7], Roberto André Kraenkel[1,2], Renato Mendes Coutinho[2,8], and Cristiana Maria Toscano[4]

[1] *São Paulo State University (UNESP), Institute for Theoretical Physics, Brazil*
[2] *Observatório Covid-19 BR*
[3] *São Paulo State University (UNESP), Infectious Diseases Department, Botucatu Medical School, Brazil*
[4] *Federal University of Goiás (UFG), Department of Ecology, Postgraduate Programme in Ecology and Evolution, Brazil*
[5] *Federal University of Goiás (UFG), Institute of Tropical Pathology and Public Health, Brazil*
[6] *Federal University of Rio Grande do Sul (UFRGS), Postgraduate Programme of Epidemiology (IPTSP), Brazil*
[7] *Universidade Federal do Rio Grande do Sul (UFRGS), Institute of Mathematics and Statistics, Department of Statistics, Brazil*
[8] *Federal University of ABC (UFABC), Center for Mathematics, Computation and Cognition, Brazil*

November 18, 2021

### 1 Introduction

In this material, we describe the methodology used in the paper. The code is available at https://github.com/covid19br/paper_markov. In section 2.1 we describe the basic epidemiological model. In section 2.2 we describe how we add vaccination to the model. In section 2.3 we describe the optimization model for vaccine doses allocation. In section 2.4 we describe how we allocate vaccine doses between ages. Finally, in section 3 we describe the parameters used in the model, together with sources. In section 4 we show the tabulated results.

### 2 Model

#### 2.1 Epidemiological model

Following Karlin and Taylor [4], we construct a discrete time Markov chain with constant probabilities. We assume that a susceptible individual ($S$) has a probability $p$ of being infected. If a infection occurs, the individual transits to the exposed ($E$), pre-symptomatic, compartment. After the incubation period, the individual can transit to hospitalized ($H$), mildly symptomatic ($I$) or asymptomatic ($A$) compartments. If the individual is hospitalized, the possible outcomes are recovery (R) or death (D), with the respective compartments. In the case of asymptomatic and mildly symptomatic we assume recovery as the only possibility.

As some parameters can be described as rates ($1/t$), we assume (as in typical SEIR-like models) an exponential distribution of time to transition. For example, the incubation period $\gamma$, can be written as the probability of a pre-symptomatic individual exiting the Exposed compartment, given by $1 - \exp(-1/\gamma)$, as follows from the Gillespie algorithm. Ignoring age structure and vaccination, the model can be written by:

$$
\begin{aligned}
S^{t+1} &= (1-p)S^t && (1)\\
E^{t+1} &= pS^t + \exp(-1/\gamma)E^t && (2)\\
I^{t+1} &= (1-\sigma)(1-\alpha)(1-\exp(-1/\gamma))E^t + \exp(-1/\nu)I^t && (3)\\
A^{t+1} &= (1-\sigma)\alpha(1-\exp(-1/\gamma))E^t + \exp(-1/\nu)A^t && (4)\\
H^{t+1} &= \sigma(1-\exp(-1/\gamma))E^t + \exp\left(-1/\nu\right)H^t && (5)\\
R^{t+1} &= R^t + (1-\mu)(1-\exp(-1/\nu))H^t + (1-\exp(-1/\nu))A^t + (1-\exp(-1/\nu))I^t && (6)\\
D^{t+1} &= D^t + \mu(1-\exp(-1/\nu))H^t && (7)
\end{aligned}
$$

Or in the form of a transition probability matrix:

$$
\mathbf{P} = \begin{pmatrix}
1-p & p & 0 & 0 & 0 & 0 & 0\\
0 & e^{-1/\gamma} & (1-\sigma)(1-\alpha)(1-e^{-1/\gamma}) & (1-\sigma)\alpha(1-e^{-1/\gamma}) & \sigma(1-e^{-1/\gamma}) & 0 & 0\\
0 & 0 & e^{-1/\nu} & 0 & 0 & 1-e^{-1/\nu} & 0\\
0 & 0 & 0 & e^{-1/\nu} & 0 & 1-e^{-1/\nu} & 0\\
0 & 0 & 0 & 0 & e^{-1/\nu} & (1-\mu)(1-e^{-1/\nu}) & \mu(1-e^{-1/\nu})\\
0 & 0 & 0 & 0 & 0 & 1 & 0\\
0 & 0 & 0 & 0 & 0 & 0 & 1
\end{pmatrix} \quad (8)
$$

where the evolution of the system is given by the Chapman-Kolmogorov equation [7]:

$$
\mathbf{x^{t+1}} = \mathbf{Px^t} \quad (9)
$$

where $\mathbf{x^t}$ is a column vector given by:

$$
\mathbf{x^t} = (S^t,\, E^t,\, I^t,\, A^t,\, H^t,\, R^t,\, D^t) \quad (10)
$$

Notice that we now can write the compartments as vector quantities describing age structure. We divide our population in 10-year age bins, thus, for example $S$, is written as $S = (S_1, S_2, \ldots, S_{10})$ and the transition matrix has the probabilities rewritten as diagonal matrices, giving different values for each age bin, if necessary (see Table S1).

#### 2.2 Vaccination Model

To account for vaccination we duplicate the model for first and second dose vaccinated individuals for each vaccine (AZD1222, CoronaVac, BNT162b2), as they are simulated at the same time. The equations for these compartments are given by:

$$
\begin{aligned}
S^{t+1}_{j,v} &= (1-p_{j,v})S^t_{j,v} && (11)\\
E^{t+1}_{j,v} &= p_{j,v}S^t_{j,v} + \exp(-1/\gamma)E^t_{j,v} && (12)\\
I^{t+1}_{j,v} &= (1-\sigma_{j,v})(1-\alpha_{j,v})(1-\exp(-1/\gamma))E^t_{j,v} + \exp(-1/\nu)I^t_{j,v} && (13)\\
A^{t+1}_{j,v} &= (1-\sigma_{j,v})\alpha_{j,v}(1-\exp(-1/\gamma))E^t_{j,v} + \exp(-1/\nu)A^t_{j,v} && (14)\\
H^{t+1}_{j,v} &= \sigma_{j,v}(1-1/\exp(-\gamma))E^t_{j,v} + \exp\left(-1/\nu\right)H^t_{j,v} && (15)\\
R^{t+1}_{j,v} &= R^t_{j,v} + (1-\mu_{j,v})(1-\exp(-\nu))H^t_{j,v} + (1-\exp(-1/\nu))A^t_{j,v} + (1-\exp(-1/\nu))I^t_{j,v} && (16)\\
D^{t+1}_{j,v} &= D^t_{j,v} + \mu_{j,v}(1-\exp(-1/\nu))H^t_{j,v} && (17)
\end{aligned}
$$

where $j$ accounts for number of doses and $v$ accounts for vaccine type. And the parameters are written as done by Ferreira et al. [2]:

$$
\begin{aligned}
p_{j,v} &= (1-\epsilon_{p,j,v})p\\
\alpha_{j,v} &= 1-(1-\epsilon_{\alpha,j,v})(1-\alpha)\\
\sigma_{j,v} &= (1-\epsilon_{\sigma,j,v})\sigma\\
\mu_{j,v} &= (1-\epsilon_{\sigma,j,w})\mu
\end{aligned} \quad (18)
$$

Notice that the protection parameters are different than efficacies, since we follow the mathematical approach to remove multiplicative effects of efficacies done in Ferreira et al. [2] (notice that our $p$ is equivalent to $\beta$ in their model).

We have now to specify how the transitions from unvaccinated to vaccinated individuals happen. Since the vaccination rates can change in time, it does not obey the assumption of exponential time distribution, therefore it is not trivial to modify the transition matrix. We then transfer the individuals to the vaccinated classes using an external vector $\mathbf{v}$, thus we have

$$\mathbf{x}^* = \mathbf{P}\mathbf{x}^{\mathbf{t}} \tag{19}$$

$$\mathbf{x}^{\mathbf{t+1}} = \mathbf{x}^* + \mathbf{v} \tag{20}$$

where $\mathbf{v}$ is vaccine and age stratified.

#### 2.3 Optimization model

What is left to us is to calculate the values of $\mathbf{v}$ in time. We follow a similar fashion of Ferreira et al. [2], where we use an optimization model to allocate doses to first and second inoculations, guaranteeing the availability of doses for the second shot, while keeping the number of doses in stock to a minimum. In this work, we generalize the work by Ferreira et al. [2] by assuming that the production/deployment rate of vaccine doses can vary in time, and also that individuals could have been vaccinated previously with the first dose.

We write a dynamical equation for the vaccine stock $V(t)$ assuming a previously defined production rate $p(t)$ and a varying withdrawal rate given by the vaccination rate, which can be chosen, that is, it is the control variable. We impose that a constant fraction $\theta' = 1 - \theta$ of the people who take the first dose will receive the second one after a period $a$, so we must be careful that the total vaccination rate is the sum of both first and second dose vaccination rates, but the control variable $v(t)$ is the vaccination rate of first doses only. We also assume that there's an initial stock of vaccines $V_0$, and an initial history of people who have already received the first dose, which show up as a rate $g(t)$ of second doses in the beginning of the roll-out (that is, for $t \in [0, a)$). The equation for $V(t)$ then is:

$$\begin{cases} \dfrac{dV}{dt} & = p(t) - v(t) - \theta' v(t-a) - g(t) \\ V(0) & = V_0 \ , \qquad v(t) = 0 \, \forall \, t < 0 \end{cases} \tag{21}$$

We note already that we can solve this equation, obtaining

$$V(t) = V_0 + \int_0^t p(t')dt' - \int_0^t v(t')dt' - \theta' \int_0^{t-a} v(t')dt' - \int_0^t g(t')dt' \tag{22}$$

We define the optimization problem by stating the objective function to be minimized and restrictions that the solution must obey. Since we want to use vaccine doses as quickly as possible, a reasonable goal is to minimize the stock of vaccines $V(t)$. With that, we impose that total vaccination rate is limited by a certain maximum value, and of course it is positive; also, the vaccine stock $V(t)$ is always positive. Finally, we must ensure that, in the period after the simulation ends ($t > T$) there will be enough doses left to apply the second doses on those who have already taken the first dose, here assuming that the production rate after the simulation interval will be the same as the value in the end of the simulation.

These considerations lead to the following optimization problem:

$$\begin{aligned} &\text{find } \min_f J = \int_0^T V(t)dt \text{ subject to} \\ &v(t) \geq 0 \\ &v(t) + \theta' v(t-a) + g(t) \leq v_{max} \\ &V(t) \geq 0 \\ &V(T) \geq \theta' \int_{T-a}^T v(t')dt' - p(T)a \end{aligned} \tag{23}$$

**Solution**

We can solve the problem defined by Eqs. (22, 23) using linear programming. This is feasible because the objective function and all constraints are linear functions of the control variable $f$ and state variable $V$ and, as Eq.(22) shows, $V$ is linear on the control variable.

We first discretize the time in $n$ intervals of length $\Delta t = \frac{T}{n}$, each interval ending at $t_i$, $i = 1, \ldots, n$, and assume that the production rate, the initial second-dose rate, and the control function will be constant over each interval (that is, a step function), with values $\vec{p} = (p(t_1), p(t_2), \ldots, p(t_n))$, $\vec{g} = (g(t_1), g(t_2), \ldots, g(t_n))$, and $\vec{x} = (v(t_1), v(t_2), \ldots, v(t_n))$, with $g(t) = 0$ when $t > a$. Eq.(22) then becomes

$$V(t_i) = V_0 + \sum_{j=1}^{i} p_i - \sum_{j=1}^{i} x_j - \theta' \sum_{j=1}^{i-\hat{a}} x_j - \sum_{j=1}^{i} g_i \ ,$$

and we seek to minimize the objective function (given by Eq.(23), up to a constant) that is a linear function of $\vec{x}$, subject to the (linear) restrictions.

The discrete version of the problem becomes:

$$\begin{aligned}
\min_{\vec{x}} J = \min_{\vec{x}} - \sum_{j=1}^{n} \left[ \sum_{i=1}^{j} x_i + \theta' \sum_{i=1}^{j-\hat{a}} x_i \right] \quad \text{subject to} \\
\vec{x} \geq 0 \\
x_i + \theta' x_{i-\hat{a}} + g_i \leq v_{max} \ , \text{ for } i = 1, \ldots, n \\
\sum_{i=1}^{j} x_i + \theta' \sum_{i=1}^{j-\hat{a}} x_i \leq V_0 + \sum_{i=1}^{j} p_i - \sum_{i=1}^{j} g_i \ , \text{ for } j = 1, \ldots, n \\
(1+\theta') \sum_{i=1}^{n} x_i \leq V_0 + \sum_{i=1}^{n} p_i + p_n a \ ,
\end{aligned} \tag{24}$$

where $\hat{a} = \frac{a}{\Delta t}$ (chosen so that $\hat{a}$ is integer), and, to simplify notation, $x_i$ is taken to be zero over values of $i$ below 1. Moreover, some care must be taken with the function $g$ for the system to be feasible: we have to guarantee that $g_j \leq v_{max}$ and $\sum_{i=1}^{j} g_i \leq V_0 + \sum_{i=1}^{j} p_i$ for $j = 1, 2 \ldots \hat{a}$.

These conditions can readily be written in matrix form and solved using standard linear programming algorithms. We implemented them in R using the package `lpSolve` [1] to solve the linear programming problem. Each vaccine is run independently.

#### 2.4 Dose allocation by age and vaccine

The solution of the optimization problem gives us the number of shots given for first and second doses by day, without differentiating between classes of the model. We consider two scenarios, the first assumes that all available shots will be used in the oldest age-bin before making vaccines available to younger ones. The second scenario assumes that vaccines will be available to younger individuals if a coverage threshold is reached by the older age bins.

##### First Scenario

For the first scenario, we have for the first dose:

$$v^t_{S,a,j,1} = \min\left(\sum_i V^t_{i,1}, S^t_a + R^t_a\right) \frac{S^t_a}{S^t_a + R^t_a} \frac{V^t_{j,1}}{\sum_i V^t_{i,1}} \tag{25}$$

$$v^t_{R,a,j,1} = \min\left(\sum_i V^t_{i,1}, S^t_a + R^t_a\right) \frac{R^t_a}{S^t_a + R^t_a} \frac{V^t_{j,1}}{\sum_i V^t_{i,1}} \tag{26}$$

where $V^t_{j,1}$ accounts for all vaccine doses of type $j$ available to first inoculation at time $t$ and $a$ gives the age bin being currently vaccinated. The first term accounts for not vaccinating more than available individuals. The first ratio accounts for proportionality between unvaccinated populations of susceptible and recovered individuals and the second ratio accounts for proportionality of number of doses in stock for each vaccine (thus we not consider rational choice of vaccine type). Notice also that we only vaccinate susceptible or recovered individuals since the epidemiologic dynamics is much faster than protection building generated by vaccination.

Second dose vaccination follows the same idea, with the difference of not having to be proportional to vaccine stock since we do not model interchangeability of vaccines, Setting $V^t_{j,a,1} = v^t_{S,a,j,1} + v^t_{R,a,j,1}$, i.e. the vaccine doses of type $j$ used for first dose in age bin $a$ at time $t$, we have:

$$v^t_{S,a,j,2} = \min\left(\theta' V^{t-\omega_j}_{j,a,1}, S^t_{a,j,1} + R^t_{a,j,1}\right) \frac{S^t_{a,j,1}}{S^t_{a,j,1} + R^t_{a,j,1}} \tag{27}$$

$$v^t_{R,a,j,2} = \min\left(\theta' V^{t-\omega_j}_{j,a,1}, S^t_{a,j,1} + R^t_{a,j,1}\right) \frac{R^t_{a,j,1}}{S^t_{a,j,1} + R^t_{a,j,1}} \tag{28}$$

where $\theta'$ accounts for the proportion of individuals that appears for second dose inoculation and $\omega_j$ is the time interval between doses assumed for vaccine $j$ (28 days for CoronaVac, for example). By using the "delayed" vaccination rate, we ensure that the same quantity of individuals that receive the first dose $\omega$ days before receive the second dose. If there is any vaccine dose available to younger ages, the procedure is repeated until total consumption of vaccine doses (or unvaccinated population). $\mathbf{v}$ is then written appropriately to transfer the individuals between compartments.

##### Second Scenario

The second scenario consider making vaccine doses available to younger ages before total coverage of older individuals. Considering a preset coverage threshold $T$, at each iteration of the model, we calculate beforehand the age classes that have coverage equal or greater than $T$. Then, the "open" age bins are one lower than the calculated and the index is given by $D$. For first dose, the distribution of doses is weighted by the number of unvaccinated individuals in each age bin. Thus, we have:

$$v^t_{S,a,j,1} = \min\left(\sum_i V^t_{i,1}, S^t_a + R^t_a\right) \frac{S^t_a}{\sum_{i=D}^{N} S^t_i + R^t_i} \frac{V^t_{j,1}}{\sum_i V^t_{i,1}} \tag{29}$$

$$v^t_{R,a,j,1} = \min\left(\sum_i V^t_{i,1}, S^t_a + R^t_a\right) \frac{R^t_a}{\sum_{i=D}^{N} S^t_i + R^t_i} \frac{V^t_{j,1}}{\sum_i V^t_{i,1}} \tag{30}$$

and for the second dose:

$$v^t_{S,a,j,2} = \min\left(\theta' V^{t-\omega_j}_{j,a,1}, S^t_{a,j,1} + R^t_{a,j,1}\right) \frac{S^t_{a,j,1}}{S^t_{a,j,1} + R^t_{a,j,1}} \tag{31}$$

$$v^t_{R,a,j,2} = \min\left(\theta' V^{t-\omega_j}_{j,a,1}, S^t_{a,j,1} + R^t_{a,j,1}\right) \frac{R^t_{a,j,1}}{S^t_{a,j,1} + R^t_{a,j,1}} \tag{32}$$

following the same idea of the first scenario. If there is any vaccine dose available to younger ages, the procedure is repeated until total consumption of vaccine doses (or unvaccinated population). $\mathbf{v}$ is then written appropriately to transfer the individuals between compartments.

#### 3 Parameterization

Table S1 comprises the parameters used in the model. Table S2 comprises the values of age dependent parameters of the model. Table S3 comprises projected vaccine quantities by the Ministry of Health (from [6]). Fig S1 illustrates the deployment rate of vaccines as function of time.

| Parameter | Description | Value | Source |
|---|---|---|---|
| $p$ | Probability of infection of susceptible individual per day | see main text | Assumed |
| age dist | Estimated age distribution of Brazil as 2020 | Table S2 | IBGE [3] |
| $\gamma$ | Incubation period of the disease | 5.8 | Wei et al. [11] |
| $\sigma$ | Infection hospitalization rate | Table S2 | Salje et al. [8] |
| $\alpha$ | Proportion of asymptomatic individuals | Table S2 | [0-20)[9]<br>[20-120)[10] |
| $\nu$ | Time to recovery/death<br>(assumed equal for any disease severity) | Table S2 | SIVEP-Gripe [5] |
| $\mu$ | Proportion of hospitalized individuals that die | Table S2 | SIVEP-Gripe [5] |

**Table S1:** Parameters used in the model with sources.

| Age bin | age dist | $\sigma$ | $\mu$ | $\alpha$ | $1-\alpha$ | $\nu$ (days) |
|---|---|---|---|---|---|---|
| [0-10) | 29,380,622 | 0.001 | 0.071 | 0.695 | 0.305 | 9.27 |
| [10-20) | 30,596,341 | 0.001 | 0.11 | 0.695 | 0.305 | 9.9 |
| [20-30) | 34,219,132 | 0.005 | 0.131 | 0.44 | 0.56 | 8.96 |
| [30-40) | 34,231,961 | 0.011 | 0.155 | 0.44 | 0.56 | 9.45 |
| [40-50) | 29,255,478 | 0.014 | 0.208 | 0.44 | 0.56 | 10.41 |
| [50-60) | 23,875,081 | 0.029 | 0.287 | 0.44 | 0.56 | 11.61 |
| [60-70) | 16,732,972 | 0.058 | 0.408 | 0.31 | 0.69 | 12.71 |
| [70-80) | 9,023,052 | 0.093 | 0.516 | 0.31 | 0.69 | 12.79 |
| [80-90) | 3,625,888 | 0.262 | 0.602 | 0.31 | 0.69 | 11.69 |
| [90-120) | 815,165 | 0.262 | 0.678 | 0.31 | 0.69 | 10.31 |

**Table S2:** Age dependent parameters separated by age bins.

| Month | AZD1222 | CoronaVac | BNT162b2 |
|---|---|---|---|
| August | 11,600,000 | 20,000,000 | 33,300,000 |
| September | 11,951,500 | 17,115,672 | 37,495,530 |
| Oct to Dec | 55,919,600 | 0 | 99,999,900 |

**Table S3:** Projected number of doses by vaccine and month.

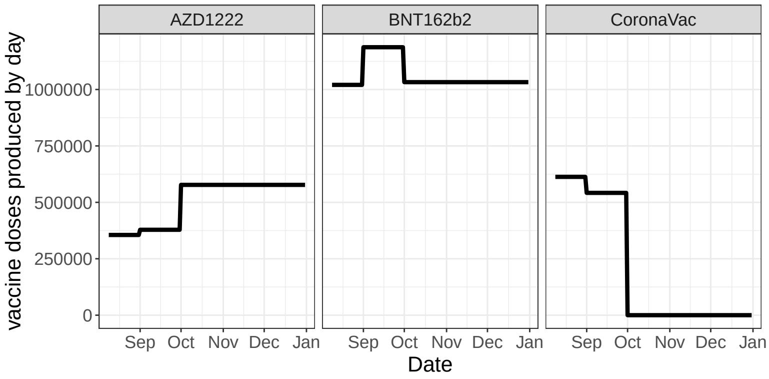


**Figure S1:** Vaccine doses projected to be produced by type of vaccine.

### 4 Results

In this section we show the results in table format. Table S4 comprises the results concerning coverage threshold and excess of deaths. Table S5 comprises the reduction in deaths by reducing interval between doses of AZD1222 in an ideal vaccination programme. Table S6 comprises the reduction in deaths by reducing interval between doses of AZD1222 with the current projected doses by the Ministry of Health (Table S3). Finally, Tables S7 and S8 comprises the results concerning the reduction in deaths caused by increase of production of AZD1222.

| Probability of infection | Coverage threshold (%) | Excess of deaths | 95% CI |
|---|---|---|---|
| Very low | 10 | 212.97 | 214.12-256.90 |
| | 20 | 212.97 | 214.12-256.90 |
| | 30 | 212.97 | 214.12-256.90 |
| | 40 | 201.04 | 202.07-242.57 |
| | 50 | 191.5 | 192.48-231.48 |
| | 60 | 167.93 | 169.00-203.32 |
| | 70 | 137.84 | 138.56-167.57 |
| | 80 | 83.42 | 84.02-102.58 |
| | 90 | 10.57 | 10.60-13.43 |
| Low | 10 | 4780.72 | 4817.59-5788.25 |
| | 20 | 4780.72 | 4817.59-5788.25 |
| | 30 | 4780.72 | 4817.59-5788.25 |
| | 40 | 4509.89 | 4544.69-5461.25 |
| | 50 | 4290.55 | 4324.54-5202.03 |
| | 60 | 3757.32 | 3787.30-4563.93 |
| | 70 | 3073.25 | 3098.22-3753.94 |
| | 80 | 1871.93 | 1881.89-2304.52 |
| | 90 | 232.05 | 232.64-291.15 |
| Medium | 10 | 8587.94 | 8665.31-10442.08 |
| | 20 | 8587.94 | 8665.31-10442.08 |
| | 30 | 8587.94 | 8665.31-10442.08 |
| | 40 | 8095.57 | 8166.75-9844.54 |
| | 50 | 7692.68 | 7765.54-9365.58 |
| | 60 | 6727.47 | 6786.80-8201.17 |
| | 70 | 5484.90 | 5518.52-6705.85 |
| | 80 | 3315.35 | 3331.87-4088.31 |
| | 90 | 400.67 | 400.72-501.53 |
| High | 10 | 14048.19 | 14105.32-17182.06 |
| | 20 | 14048.19 | 14105.32-17182.06 |
| | 30 | 14048.19 | 14105.32-17182.06 |
| | 40 | 13225.35 | 13276.29-16176.70 |
| | 50 | 12542.13 | 12589.76-15366.06 |
| | 60 | 10945.61 | 10985.07-13408.18 |
| | 70 | 8883.86 | 8917.38-10906.72 |
| | 80 | 5306.95 | 5325.87-6542.27 |
| | 90 | 620.21 | 617.35-774.56 |

**Table S4:** Excess of deaths caused by premature vaccination of younger individuals. Results appear in Fig. 1 of the main text.

| Probability of infection | Interval between doses (weeks) | Reduction in deaths | 95% CI |
|---|---|---|---|
| Very Low | 8 | 99.56 | 64.17-133.82 |
| | 9 | 79.34 | 50.40-107.54 |
| | 10 | 55.83 | 35.23-76.23 |
| | 11 | 32.96 | 21.31-44.55 |
| Low | 8 | 2451.48 | 1598.06-3273.52 |
| | 9 | 1947.00 | 1255.87-2620.37 |
| | 10 | 1355.21 | 854.36-1837.43 |
| | 11 | 781.16 | 502.75-1050.65 |
| Medium | 8 | 4799.96 | 3155.86-6398.54 |
| | 9 | 3806.35 | 2493.27-5103.71 |
| | 10 | 2621.67 | 1658.09-3530.17 |
| | 11 | 1478.42 | 943.62-1987.47 |
| High | 8 | 9146.34 | 5941.56-12298.09 |
| | 9 | 7211.36 | 4595.12-9817.33 |
| | 10 | 4887.34 | 3015.67-6712.7 |
| | 11 | 2655.41 | 1665.4-3602.38 |

**Table S5:** Reduction in deaths caused by reducing interval between doses of AZD1222 without limitation of number of doses. Results appear in Fig. 2 of the main text.

| Probability of infection | Interval between doses (weeks) | Reduction in deaths | 95% CI |
|---|---|---|---|
| Very Low | 8 | 0.00 | 0.00-0.01 |
| | 9 | 0.00 | 0.00-0.00 |
| | 10 | 0.00 | 0.00-0.00 |
| | 11 | 0.00 | 0.00-0.00 |
| Low | 8 | 0.09 | 0.02-0.16 |
| | 9 | 0.07 | 0.00-0.13 |
| | 10 | 0.07 | 0.00-0.13 |
| | 11 | 0.07 | 0.00-0.13 |
| Medium | 8 | 0.19 | 0.05-0.31 |
| | 9 | 0.16 | 0.03-0.26 |
| | 10 | 0.16 | 0.03-0.26 |
| | 11 | 0.16 | 0.03-0.26 |
| High | 8 | 0.38 | 0.13-0.58 |
| | 9 | 0.34 | 0.11-0.53 |
| | 10 | 0.34 | 0.11-0.53 |
| | 11 | 0.34 | 0.11-0.53 |

**Table S6:** Reduction in deaths caused by reducing interval between doses of AZD1222 with the current number of projected doses. Results appear in Fig. 3 of the main text.

| Probability of infection | Interval between doses (weeks) | Increase in production (%) | Reduction in deaths | 95% CI |
|---|---|---|---|---|
| Very Low | 8 | 0 | 0.01 | 0.00-0.01 |
| | | 25 | 0.82 | 0.36-1.22 |
| | | 50 | 4.36 | 1.84-6.51 |
| | | 75 | 13.61 | 7.45-19.40 |
| | | 100 | 20.56 | 11.3-29.03 |
| | 9 | 0 | 0.01 | 0.00-0.01 |
| | | 25 | 0.74 | 0.32-1.10 |
| | | 50 | 4.10 | 1.70-6.14 |
| | | 75 | 13.14 | 7.20-18.74 |
| | | 100 | 19.77 | 10.88-27.90 |
| | 10 | 0 | 0.01 | 0.00-0.01 |
| | | 25 | 0.71 | 0.31-1.07 |
| | | 50 | 3.90 | 1.62-5.84 |
| | | 75 | 12.52 | 6.97-17.75 |
| | | 100 | 18.53 | 10.33-26.03 |
| | 11 | 0 | 0.01 | 0.00-0.01 |
| | | 25 | 0.50 | 0.14-0.79 |
| | | 50 | 3.12 | 1.25-4.72 |
| | | 75 | 11.03 | 6.44-15.38 |
| | | 100 | 14.56 | 8.25-20.38 |
| Low | 8 | 0 | 0.13 | 0.06-0.19 |
| | | 25 | 16.64 | 6.25-25.70 |
| | | 50 | 105.82 | 44.36-157.39 |
| | | 75 | 310.59 | 169.15-441.57 |
| | | 100 | 496.11 | 280.81-703.06 |
| | 9 | 0 | 0.13 | 0.06-0.19 |
| | | 25 | 15.07 | 5.41-23.50 |
| | | 50 | 100.50 | 41.56-149.98 |
| | | 75 | 300.63 | 163.88-427.57 |
| | | 100 | 478.02 | 268.29-675.22 |
| | 10 | 0 | 0.13 | 0.06-0.19 |
| | | 25 | 14.59 | 5.17-22.83 |
| | | 50 | 95.94 | 39.55-143.26 |
| | | 75 | 285.32 | 156.96-404.34 |
| | | 100 | 444.13 | 248.71-622.53 |
| | 11 | 0 | 0.13 | 0.06-0.19 |
| | | 25 | 13.77 | 5.27-21.18 |
| | | 50 | 79.50 | 32.53-118.85 |
| | | 75 | 248.32 | 144.05-347.72 |
| | | 100 | 376.10 | 224.31-518.26 |

**Table S7:** Reduction in deaths caused by reducing interval between doses of AZD1222 and increasing the number of doses produced, for very low and low probabilities of infection. Results appear in Fig. 4 of the main text.

| Probability of infection | Interval between doses (weeks) | Increase in production (%) | Reduction in deaths | 95% CI |
|---|---|---|---|---|
| Medium | 8 | 0 | 0.25 | 0.11-0.36 |
| | | 25 | 38.44 | 17.72-56.41 |
| | | 50 | 205.05 | 86.60-301.85 |
| | | 75 | 620.25 | 341.31-882.8 |
| | | 100 | 1009.33 | 587.16-1429.56 |
| | 9 | 0 | 0.24 | 0.11-0.35 |
| | | 25 | 36.13 | 16.56-53.18 |
| | | 50 | 196.94 | 82.00-290.75 |
| | | 75 | 604.18 | 332.31-861.06 |
| | | 100 | 975.44 | 568.78-1370.74 |
| | 10 | 0 | 0.24 | 0.11-0.35 |
| | | 25 | 35.40 | 16.18-52.14 |
| | | 50 | 186.54 | 76.73-271.63 |
| | | 75 | 574.96 | 320.94-817.50 |
| | | 100 | 911.22 | 533.01-1278.87 |
| | 11 | 0 | 0.24 | 0.11-0.35 |
| | | 25 | 33.23 | 15.40-48.43 |
| | | 50 | 157.86 | 65.45-233.44 |
| | | 75 | 500.26 | 294.94-702.29 |
| | | 100 | 769.53 | 460.20-1063.99 |
| High | 8 | 0 | 0.43 | 0.18-0.63 |
| | | 25 | 67.91 | 33.96-98.89 |
| | | 50 | 375.75 | 158.83-551.02 |
| | | 75 | 1213.50 | 677.49-1723.24 |
| | | 100 | 2018.58 | 1167.27-2823.14 |
| | 9 | 0 | 0.43 | 0.18-0.62 |
| | | 25 | 65.36 | 32.86-95.43 |
| | | 50 | 366.36 | 154.68-538.26 |
| | | 75 | 1191.99 | 666.74-1691.30 |
| | | 100 | 1966.14 | 1138.37-2746.82 |
| | 10 | 0 | 0.43 | 0.18-0.62 |
| | | 25 | 64.47 | 32.48-94.18 |
| | | 50 | 353.23 | 149.18-521.67 |
| | | 75 | 1133.36 | 637.63-1602.48 |
| | | 100 | 1847.51 | 1089.35-2565.96 |
| | 11 | 0 | 0.43 | 0.18-0.62 |
| | | 25 | 58.81 | 29.94-85.16 |
| | | 50 | 304.72 | 132.46-445.86 |
| | | 75 | 991.74 | 579.41-1379.72 |
| | | 100 | 1566.21 | 931.57-2142.29 |

**Table S8:** Reduction in deaths caused by reducing interval between doses of AZD1222 and increasing the number of doses produced, for medium and high probabilities of infection. Results appear in Fig. 4 of the main text.
